# Measuring Total Healthcare Utilization Among Intimate Partner Violence Survivors In Primary Care

**DOI:** 10.1101/19007377

**Authors:** Mary E. Logeais, Qi Wang, Lynette M. Renner, Cari Jo Clark

## Abstract

Rising health care costs are influenced by health care utilization, which encompasses hospital, ambulatory and non-face-to-face episodes of care. In this study, we created a novel a health care utilization-scoring tool that was used to examine whether one psychosocial factor, intimate partner violence (IPV), leads to higher utilization of health care services when controlling for relevant confounders. We sought to fill gaps about how social and behavioral issues impact utilization—particularly non-face-to-face episodes of care.

We conducted a retrospective cross-sectional study in 2017 examining patients seen at 11 University-affiliated primary care clinics from January 2015 to December 2016 who were screened for IPV. A total of 31,305 patients were screened, of which 280 screened positive. We controlled for medical complexity by deriving the revised Charlson Comorbidity Index for each patient. We calculated a novel utilization score, which was a weighted sum of hospital, ambulatory and non-face-to-face encounters. Missed appointments were also measured.

IPV-positive and IPV-negative patients were similar with respect medical complexity. IPV-positive patients had significantly higher mean utilization scores (54 vs. 40, *p*<0.001) and more missed appointments (3 vs. 1.3, *p*<0.001). IPV was associated with increased total utilization (*p*=0.015), as well as non-face-to-face and ambulatory visits (*p*=0.025 and *p*=0.015, respectively) for female patients and was associated with more missed appointments for both males and females (*p*< .001).

These data support more inclusive population-specific interventions focusing on social determinants of health to reduce both face-to-face and non-face-to-face utilization, which may improve health care expenditures, outcomes and provider satisfaction.

## 1. INTRODUCTION

In the United States, a disproportionate share of health care spending is attributed to a relatively small group of patients. Thus, as national health care costs continue to rise, health care utilization is a critical research focus. The Agency for Healthcare Research and Quality estimates the top 5% of patients account for approximately 50% of U.S. health care expenditures, while the top 1% of patients account for 21% of healthcare costs (Cohen and Yu 2012). Health care utilization is most often analyzed within the economic context of volumes of services rendered (Da Silva et al. 2011), with the term ‘high-utilizer’ applied to patients who accumulate large numbers of emergency department visits or hospital admissions (Mann 2013). However, health care utilization should be expanded to include a wide range of ambulatory services and non-face-to-face episodes of care, whereby a patient may interact with the health system outside of a traditional office visit. Although it is not customary to bill for non-face-to-face episodes under a traditional fee-for-service model of care, these encounters may require significant provider time or clinic resources (Christino et al. 2013). Non-face-to-face encounters include telephone calls, prescription refills, email messages, or review of reports or records (Baron 2010). The authors of one observational analysis of ambulatory activities across four specialties found that physicians spent only 33.1% of their time on direct clinical face time with patients and staff (Sinsky et al. 2016). The consequences of this allocation of effort ranged from lost revenue (an estimated $4–6 for every minute spent), to poorer patient-physician interactions, worse patient health outcomes, and greater physician burnout (Gottschalk and Flocke 2005; Shanafelt et al. 2012).

A potential solution to the utilization and payment incentive crisis requires us to identify high-utilizers of care in order to further target gaps in our care delivery system and deliver interventions to those at highest risk. High-utilizers often lack access to regular primary care, are of lower socioeconomic status, have complex medical conditions, and face challenging psychosocial factors, such as trauma, homelessness, substance abuse or mental illness (Bell et al. 2017; Mann 2013). Additionally, females account for a larger overall burden of health care costs compared to males, owing in part to reproductive health needs and longer lifespans (Owens 2008). One additional psychosocial factor associated with more frequent utilization of health care services is intimate partner violence (IPV). IPV, defined as physical violence, sexual violence, stalking and/or psychological aggression by a current or former intimate partner, affects an estimated one in three females and one in four males in their lifetime and leads to numerous adverse physical, mental and behavioral health outcomes (Breiding et al. 2014; Breiding, Chen, and Black 2014; Breiding, Black, and Ryan 2008). IPV is associated with increased utilization of emergency care and other services, even after violence has stopped (Fishman et al. 2010; Jones et al. 2006). Females who experienced IPV in the past year had health care costs that were 122% higher than females without a history of IPV, and 60% higher than females who experienced IPV prior to the past year (Bonomi et al. 2009; Rivara et al 2007). Using data from the Medical Expenditure Panel Survey and the National Violence Against Women Survey, the estimated costs of IPV at the U.S. population level are $2.6 billion per year (Brown, Finkelstein, and Mercy 2008; Max et al. 2004). Due to the prevalence of IPV victimization and its exorbitant costs, understanding how IPV is associated with increased health care utilization is the first step in disrupting this association.

In this study, we built upon existing IPV and utilization literature by expanding the scope of health care utilization examined. To date, there are no published findings specific to IPV or chronic disease where researchers examined non-face-to-face encounters when assessing utilization— despite the growing frequency, associated costs, and professional dissatisfaction with these encounters (Blumenthal et al. 2017; Heincelman et al. 2016; Meijs, et al. 2014; Robinson et al. 2014; Seibert et al. 2008). To fill this gap, we developed a novel health care utilization scoring tool—which encompasses both face-to-face encounters and non-face-face encounters—to assess whether patients who experience IPV have a higher utilization of health care services compared to those who do not when controlling for relevant confounders. Through this study, we sought to shed light on how IPV victimization is associated with differences in utilization for females and males; and consequently, how population-specific interventions may improve health care expenditures, outcomes and provider satisfaction, satisfying the ‘quadruple aim’ of health care (Bodenheimer and Sinsky 2014).

## 2. METHODS

### 2.1 Setting

Our study involved patients seen within primary care clinics associated with a large Midwest university. The university hospital and clinics serve as a major learning site for students in the health professions as well as a site for clinically oriented research. The physician practice is subspecialty driven, consisting of approximately 1000 physicians, of which 10-15% are in primary care. The clinics serve 2500 outpatients daily. Although most patients reside in the metropolitan area, some travel from across the state and regionally for quaternary referrals. Patients come from diverse ethnic and socioeconomic backgrounds and disproportionately represent local Somali, Hmong and Middle-Eastern immigrants.

### 2.2 Sample

The study population included adult patients over the age of 18 years seen at a primary care clinic (11 affiliated family medicine, internal medicine and womens health clinics) at least once between January 1, 2015 and December 31, 2016. Patients in the sample were screened for IPV in the clinical setting, as documented in the electronic medical record.

### 2.3 Data Source

Screening and health care utilization data were extracted retrospectively in 2017 from an electronic health record (Epic^™^) data repository, which is updated daily and contains more than 2 million patient records spanning the entire health network. Health informatics specialists performed the data extraction. The University’s Institutional Review Board deemed this study exempt.

### 2.4 Measures

#### 2.4.1 Intimate partner violence

The two items used to identify IPV victimization were embedded within a three-item home safety-screening tool. We considered a negative response to the question, “Are you safe in your relationship,” or a positive response to the question “Have there been threats or direct abuse against you or your children” as proxy indicators of IPV.

#### 2.4.2 Health care utilization

Health care utilization was quantified in terms of unique encounters in the electronic medical record (EMR) (Table 1). We also measured the number of no-show or missed appointments recorded in the EMR. Because of the multitude of different encounter types in the EMR, we conducted preliminary analyses to explore the overall frequency of various encounter types in order to determine which types were clinically relevant. We assigned each type of encounter a unique weight in order to generate a numeric summed score of each patient’s total health care utilization. Higher acuity (in-patient versus out-patient) and thus, higher cost, encounters were assigned higher weights on a three-point scale (Table 1). We addressed outliers by excluding patients who had a volume of encounters beyond four standard deviations from the mean, which aligns with approaches utilized in prior analyses (AHRQ 2017; Coker et al. 2004).

**Table 1.** Utilization Score Components.

| Table 1. Utilization Score Components |  |
| --- | --- |
| Encounter Type From Most to Least Frequent | Scoring Weight |
| *Face-to-Face* |  |
| Hospital Encounter/Hospital; Emergency Room; Surgery | 3 |
| Office Visit; Therapy Visit; Allied Health/Nurse Visit; Prenatal Office Visit; Infusion Therapy Visit; Oncology; Consultation; Care Coordination; Outpatient Visit; Behavioral/Emergency Health Treatment Plan | 2 |
| *Non-Face-to-Face* |  |
| Telephone; Refill; Orders Only; Results Only; MyChart™ Medical Advice; Hospital Laboratory; Radiology Appointment; MyChart™ Refill; Ancillary Orders; Hospital Radiology; Virtual Visit; E-Visit; Team Conference | 1 |

#### 2.4.3 Risk adjustment for comorbid conditions

We calculated the revised Charlson Comorbidity Index in order to adjust for comorbid conditions that might impact health care utilization but are not related to IPV (Quan et al. 2011). Data elements used to calculate the score included ICD-10 diagnostic codes for 12 specific conditions documented in the medical record problem list or billing data. Conditions were weighted and a score was calculated by summing across the weights (Table 2). A combined Charlson Age Index was calculated by adding a value for age (where one point was assigned for every decade above 50 years old, up to five points for age 90 or greater). Both increasing age and medical comorbidity have been shown to correlate with morbidity and mortality, and consequently, health care utilization (Charlson et al. 1994).

**Table 2.** Charlson Index Scoring.

| Table 2. Charlson Index Scoring |  |
| --- | --- |
| Age Variables | Points |
| 18-49 years old | 0 |
| 50-59 years old | 1 |
| 60-69 | 2 |
| 70-79 | 3 |
| 80-89 | 4 |
| >= 90 years old | 5 |
| Comorbidity Variables | Points |
| Chronic Pulmonary Disease; Rheumatologic Disease; Diabetes with Chronic Complications; Renal Disease | 1 |
| Congestive Heart Failure; Dementia; Mild Liver disease; Hemiplegia or | 2 |
| Paraplegia; Any Malignancy, including Leukemia and Lymphoma |  |
| Moderate or Severe Liver Disease; HIV/AIDS | 4 |
| Metastatic Solid Tumor | 6 |

### 2.5 Statistical Analysis

Descriptive statistics were calculated to examine the number of patients seen and the percent of patients who screened positive for IPV victimization. Differences in study variables (sex, race, ethnicity, age, Charlson Comorbidity Age Index, missed appointments, counts of encounters by weight, and total utilization score) by whether the patient screened positive for IPV were assessed with chi-square tests, two-sample t-tests, or Wilcoxon tests depending on the nature of the variable. A multivariate linear regression model was constructed to examine whether patients who screened positive for IPV had a higher total utilization score compared to patients who never screened positive for IPV controlling for variables which are well established correlates of IPV and healthcare utilization including: sex, race and ethnicity (non-Hispanic White, all others or missing), age, and Charlson Comorbidity Index. As the covariates are theoretically relevant to the analysis, they were retained in the multivariate model regardless of their significance. An interaction between IPV and sex was examined and was dropped from the model if it was statistically insignificant. R-square was calculated to assess the fit of each model. Similar models were constructed with the outcome being total missed appointments and counts of encounters by weight. Analyses were performed using Statistical Analysis Software (version 9.3. SAS Institute Inc., Cary, NC). A two-sided *p*-value < 0.05 was considered statistically significant.

## 3. RESULTS

During the time period of interest, 31,305 unique patients were screened for IPV victimization (Table 3). The positive screening rate was 0.9% (*n*=280). Of the 280 patients who screened positive, 79.6% were female and 20.4% were male. The mean age of the sample was 45.2 years old and 49.0% of patients were non-Hispanic White, which is less than the census-level reported data for the metro region, suggesting that patients screened at the primary care clinics in this study may represent a more diverse segment of the population. The mean Charlson Comorbidity Index among patients who screened positive for IPV was 0.86 versus 0.87 for those who screened negative (*p*=0.97), with a combined Charlson Age Score of 1.50 and 1.64 (*p*=0.30), respectively. This indicated that most patients had one chronic health condition and that the two groups did not differ significantly with respect to comorbid illness.

**Table 3.**
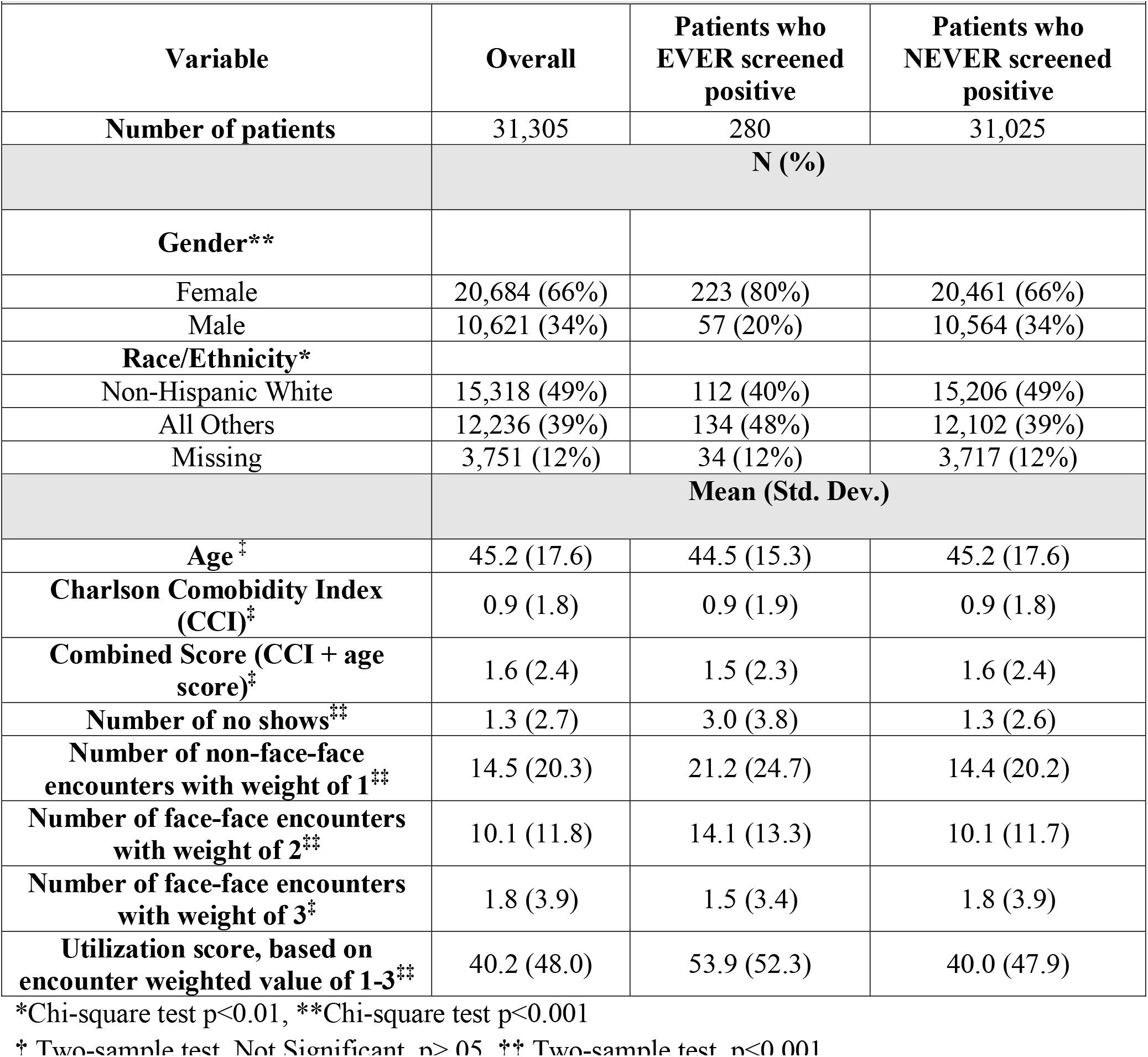
Baseline Descriptive Characteristics.

Patients who experienced IPV had a significantly higher number of missed appointments compared to patients who did not experience IPV (3.0 versus 1.3 respectively, *p*<0.001). Similarly, patients who experienced IPV had more overall health care utilization. IPV-positive patients had an average of 7 more non-face-to-face encounters and 4 additional outpatient face-to-face encounters over the two-year analysis period than IPV-negative patients (*p*<0.001). In-patient encounters (hospital, emergency department, and surgeries) were not significantly different for the two groups (1.5 versus 1.8, *p*=0.15). Despite this, the overall weighted utilization score was 54 for IPV-positive patients and 40 for IPV-negative patients (range 2-369, *p*<0.001).

In the multivariate regression analyses (Table 4), we found total health care utilization was associated with a higher Charlson Comorbidity score (*p*<0.001) and older age (*p*<0.001). Patients who were not non-Hispanic White were less likely to utilize care than non-Hispanic Whites (*p*<0.001). The main effect of IPV was not significant (*p*=0.7); however, the interaction between IPV and sex was significant (*p*=0.015). Females who experienced IPV were significantly more likely to utilize health care resources than both females who did not experience IPV (*p*<0.001) and males, with or without IPV experiences (*p*=0.004 and *p*<0.001 respectively). IPV-positive males were not more likely to utilize health care resources than IPV-negative males.

**Table 4.** Linear Regression Outcomes.

| Table 4. Linear Regression Outcomes |  |  |  |  |  |  |  |  |  |
| --- | --- | --- | --- | --- | --- | --- | --- | --- | --- |
| *Independent Vars.* | Predicting Utilization Score |  |  | Predicting Non-Face-to-Face Encounters |  |  | Predicting Missed Appointments |  |  |
|  | *Estimate* |  | *Std. Err.* | *Estimate* |  | *Std. Err.* | *Estimate* |  | *Std. Err.* |
| Female (Male Ref) | 9.26 | ** | 0.49 | 3.14 | ** | 0.20 | 0.27 | ** | 0.03 |
| All Others (Non-Hispanic White Ref) | -8.79 | ** | 0.50 | -4.71 | ** | 0.21 | 1.06 | ** | 0.03 |
| Missing (Non-Hispanic White Ref) | -9.80 | ** | 0.75 | -4.08 | ** | 0.31 | 0.53 | ** | 0.05 |
| Age | 0.34 | ** | 0.01 | 0.21 | ** | 0.01 | -0.01 | ** | 0.01 |
| Charlson Comorbidity Index | 12.29 | ** | 0.14 | 5.05 | ** | 0.06 | 0.38 | ** | 0.01 |
| IPV Positive (Negative Ref) | 2.07 | NS | 5.37 | 2.56 | NS | 2.22 | 1.55 | ** | 0.15 |
| Sex*IPV Positive | 14.61 | * | 6.02 | 5.56 | * | 2.49 |  | NS<sup>†</sup> |  |
| Intercept | 12.52 | ** | 0.83 | 0.75 | * | 0.34 | 0.58 | ** | 0.05 |
| R<sup>2</sup> | 0.29 |  |  | 0.32 |  |  | 0.09 |  |  |
| N | 31,305 |  |  | 31,305 |  |  | 31,305 |  |  |

\* $p < 0.05$ , \*\* $p < 0.0001$
†The interaction was not significant and was dropped from the final model

When examining different types of health care utilization, similar patterns held true for both non-face-to-face encounters and ambulatory visits, with IPV-positive females utilizing more resources. However, for in-patient encounters (e.g., hospital admissions, emergency room visits and surgeries), we found that neither the main effect of IPV (*p*=0.16), nor the interaction between IPV and sex was significant (*p*=0.37). Being female, non-Hispanic White, of younger age, and having a higher Charlson Comorbidity score were significantly associated with greater numbers of hospital encounters. Being female, not identifying as non-Hispanic White, being younger, having a higher Charlson Comorbidity score, and experiencing IPV victimization (females and males) (*p*<0.001) were significantly associated with missed appointments.

In summary, female patients had significantly greater total health care utilization, non-face-to-face encounters, and missed appointments (*p*<0.001). The main effect of IPV was associated with increased rates of missed appointments (*p*<0.001) and the interaction between sex and IPV was associated with increased total utilization (*p*=0.015), as well as non-face-to-face and ambulatory visits for females (*p*=0.025 and *p*=0.015, respectively).

## 4. DISCUSSION

To our knowledge, we are the first to create a health care utilization tool that includes non-face-to-face encounters and stratifies encounters by acuity (and effectively cost) in order to create a summed score. We investigated both traditional predictors of utilization, including race/ethnicity, sex, age, and medical complexity, as well as novel variables such as IPV victimization. We also examined the effects of IPV for both males and females.

Overall, the results suggest that females who experience IPV contribute higher costs to the health care system via higher utilization of care and more missed appointments than females who do not report a history of IPV. Previous studies have identified higher health care costs (Bonomi et al. 2009; Rivara et al. 2007) as well as poor utilization of prenatal care among female IPV survivors (Cha and Masho 2014). This study expands upon this literature by further characterizing associations between IPV and various types of health care encounters, including ambulatory and non-face-to-face care. We also looked at associations between IPV, utilization and missed appointments across the complete spectrum of medical and surgical specialties, rather than focusing only on women’s health or mental health visits (Cha and Masho 2014; Illangasekare S et al. 2012).

Additionally, although we found that female but not male survivors of IPV utilized more health care services, both males and females had higher numbers of missed appointments compared to patients without a reported history of IPV. This suggests that the instability that characterizes some survivors’ lives similarly influences females’ and males’ abilities to keep scheduled appointments, although it does not necessarily drive both equally towards utilization of health resources. These associations contribute negatively to costs of care and health outcomes, and support a more expansive approach to IPV screening or identification than the current clinical recommendation which focuses only on women of reproductive age (Moyer et al 2013; IOM 2011).

Of note, the IPV victimization detection rate in our study was lower than previously reported estimates of prevalence (Breiding et al. 2014) and the IPV-positive cohort included a larger than anticipated proportion of males. It is important to be aware, however, that the lower detection rate was generated through the use of two safety-focused questions that are not IPV-specific nor are a valid IPV screen (Moyer 2013). The clinic system has since adopted a 4-item validated IPV screen for use with all adult patients. It would be valuable to replicate the current study to compare rates of IPV among patients, in addition to further testing the utilization score in an effort to ensure robust functioning.

### 4.1 Limitations

This was a cross-sectional study, limiting conclusions on causality. The study was performed in a closed, albeit large, health care system using EMR data rather than claims data; thus, we did not have access to potential sources of health care utilization external to our care network. However, the study was limited to individuals with primary care visits within the system, improving the likelihood that other visit types within the system would also be captured as part of the medical home. We were limited to the inclusion of pre-existing safety items, which included proxies for IPV but did not represent a validated IPV screening tool. Approximately 12% of patients had no data on race or ethnicity captured in the EMR. We also faced challenges with respect to the clarity and accuracy of encounter-level data within the EMR, which do not consistently correlate with discrete episodes of care depending on how they are inputted or captured in the EMR, and resulted in a wide range of hospital encounter data. Researchers performing future analyses could validate hospital encounter data using discharge CPT codes or time stamp data from the EMR.

Despite these limitations, the application of the tool to assess the impact of IPV on health utilization is appropriate given prior literature; and, the utilization score developed in our study could effectively be applied to a range of patient populations and disease conditions. Our study advances the ongoing effort to identify populations of patients who disproportionately utilize health care and would benefit from early targeted interventions and enhanced care coordination. Ongoing research efforts will prove instrumental as health care reform continues to evolve to focus more on satisfying the quadruple aim, which is to deliver better quality care to populations, at lower costs, with improved provider satisfaction (Bodenheimer and Sinsky 2014).

## 5. CONCLUSION

Our results highlight an opportunity to intervene to impact both health outcomes and expenditures, particularly with respect to females and individuals who identify as racial and ethnic minorities who have experienced IPV and are utilizing more care. This is in addition to males and individuals who identify as sexual and gender minorities who may be under-recognized by health care professionals. The Institute of Medicine recommends including documentation of IPV and other social determinants of health in the electronic health record (Institute of Medicine 2014). Our study findings provide evidence to support a more coordinated and inclusive effort to identify and support patients with counseling, resources and referrals in our health care system (Institute of Medicine 2011). Additionally, while many health care organizations and payers are creating complex risk assessment models to evaluate costs and manage populations, this utilization measure has the benefit of being a comprehensive sum of health services utilization, which requires only EMR data and is replicable by scholars and clinicians.

## Data Availability

Data is housed in a data shelter and is not immediately available but can be accessed by the research team.

## ACKNOWLEDGEMENTS

We thank Hannah Scott BS and Justin Buszin PhD for their contributions to this manuscript.

This research did not receive any specific grant from funding agencies in the public, commercial or not-for-profit sectors.

No authors report any conflicts of interest or financial disclosures.

